# Epigenetic field cancerization in breast cancer using subject-matched tumor, ipsilateral-normal, and contralateral-normal tissues

**DOI:** 10.1101/19002014

**Authors:** Meghan E. Muse, Alexander J. Titus, Lucas A. Salas, Owen M. Wilkins, Chelsey Mullen, Kelly J. Gregory, Sallie S. Schneider, Giovanna M. Crisi, Rahul M. Jarwale, Christopher N. Otis, Brock C. Christensen, Kathleen F. Arcaro

**Affiliations:** Department of Epidemiology, Dartmouth Geisel School of Medicine, Hanover, NH; Department of Veterinary & Animal Sciences, University of Massachusetts, Amherst, MA; Pioneer Valley Life Sciences Institute, Baystate Medical Center, Springfield, MA; Department of Pathology, University of Massachusetts Medical School-Baystate Health, MA; Department of Molecular & Systems Biology, Dartmouth Geisel School of Medicine, Hanover, NH; Department of Community & Family Medicine, Dartmouth Geisel School of Medicine, Hanover, NH

**Author notes:** Corresponding Author: Kathleen F. Arcaro, Life Science Laboratory 1 Room N529, 240 Thatcher Road, Amherst, MA 01003-9298. Authors contributed equally (Brock C. Christensen, Kathleen F. Arcaro).

**Keywords:** Field cancerization, DNA methylation, Epigenetics, Breast cancer, Normal breast, Contralateral breast

## Abstract

**Background:** Emerging work has demonstrated that histologically normal (non-tumor) tissue adjacent to breast tumor tissue shows evidence of molecular alterations related to tumorigenesis, referred to as field cancerization effects. Although changes in DNA methylation are known to occur early in breast carcinogenesis and the landscape of breast tumor DNA methylation is profoundly altered compared with normal tissue, there have been limited efforts to identify DNA methylation field cancerization effects in histologically normal breast tissue adjacent to tumor.

**Methods:** Matched tumor, histologically normal tissue of the ipsilateral breast (ipsilateral-normal), and histologically normal tissue of the contralateral breast (contralateral-normal) were obtained from nine women undergoing bilateral mastectomy. Laser capture microdissection was used to select breast epithelial cells from normal tissues, and neoplastic cells from tumor specimens for genome-scale measures of DNA methylation with the Illumina HumanMethylationEPIC array.

**Results:** We identified substantially more CpG loci that were differentially methylated between contralateral-normal breast and tumor tissue (63,271 CpG loci *q* < 0.01), than between ipsilateral-normal tissue and tumor (38,346 CpG loci *q* < 0.01). In addition, we identified differential methylation in ipsilateral-normal relative to contralateral-normal tissue (9,562 CpG loci *p* < 0.01). Hypomethylated loci in ipsilateral normal relative to contralateral were significantly enriched for breast cancer-relevant transcription factor binding sites including those for ESR1, FoxA1, and GATA3. Hypermethylated loci in ipsilateral-normal relative to contralateral-normal tissue were significantly enriched for CpG island shore regions.

**Conclusions:** Our results indicate that early hypermethylation events in breast carcinogenesis are more likely to occur in the regions immediately surrounding CpG islands than CpG islands *per se*, reflecting a field effect of the tumor on surrounding histologically normal tissue. This work offers an opportunity to focus investigations of early DNA methylation alterations in breast carcinogenesis and potentially develop epigenetic biomarkers of disease risk.

## Background

Breast cancer is the most diagnosed non-keratinocyte cancer and is responsible for the second highest number of cancer-related deaths among women in the United States, with over 40,000 women estimated to die from breast cancer in 2019 in the United States alone [1]. It has been shown that the breast methylome varies greatly between histologically normal tissue and tumor tissue [2], and such variation is related in part to age, parity, and alcohol consumption, among other factors [3,4]. Work assessing the methylome of ductal carcinoma *in situ* (DCIS) has demonstrated that alterations to DNA methylation occur early in breast carcinogenesis [5,6]. Due to the invasive nature of sample collection, molecular studies are conducted as a comparison between tumor and histologically normal tissue adjacent to the tumor (subsequently referred to as adjacent normal) tissue. However, given that alterations to DNA methylation have been identified at the early stages of breast carcinogenesis, it is currently unknown whether adjacent normal tissue is reflective of normal breast epithelium at the molecular level. This limitation may lead to an incomplete understanding of tumor development or an attenuation of the measured magnitude of epigenetic differences.

If a field effect of the tumor on adjacent histologically normal tissue occurs in breast cancer, the use of adjacent normal tissue as a referent sample in molecular studies would inhibit our ability to identify the earliest molecular alterations that occur in carcinogenesis. Therefore, assessment of possible field cancerization effects in breast cancer is critical to informing our understanding of early carcinogenesis. A recent study demonstrated evidence of alterations to DNA methylation related to tumorigenesis in histologically normal tissue adjacent to breast tumors, suggestive of a field cancerization effect in breast cancer [7]. However, this study was conducted comparing adjacent normal tissue in women with breast cancer to normal breast tissue in women without breast cancer. While comparing cancer-free control tissue from one set of study participants with adjacent normal tissue from an independent set of participants provides initial evidence of field cancerization effects, it may obscure the extent to which DNA methylation profiles reflect individual differences/exposures and thereby modify the field effect profile. To our knowledge, no study to-date has shown evidence of field effects in breast cancer using subject-matched tumor tissue, histologically normal tissue from the ipsilateral breast (ipsilateral-normal), and histologically normal tissue from the contralateral breast (contralateral-normal). Furthermore, most prior research assessing alterations to DNA methylation in breast tumors has leveraged bulk tissue samples which may confound results due to underlying differences in the cellular composition of collected tissue. In the present study, we present evidence of field effects in breast cancer, using subject-matched, laser capture microdissected tumor, ipsilateral-normal, and contralateral-normal samples from nine women undergoing bilateral mastectomy.

## Methods

### Study population

This was a retrospective study using formalin-fixed, paraffin-embedded (FFPE) archival tissue blocks from treatment-naïve women who had undergone bilateral mastectomies (tumor mastectomy and prophylactic contralateral mastectomy) at Baystate Medical Center, Springfield, MA and who were enrolled in the Rays of Hope Center for Breast Research Registry (IRB Baystate Health, Springfield, MA protocol number 568088).

### Laser-capture microdissection and DNA isolation

Breast tumor, ipsilateral-normal, and contralateral-normal tissue blocks were identified and pulled for each of 9 women. H&E stained sections of the tumor and benign samples were used to estimate the total area for microdissection; a minimum of 5 × 10^6^ μm^2^ was required to ensure sufficient material for adequate DNA yield (∼800 ng minimum). There was insufficient tumor tissue available for one woman. Consecutive tissue sections (6-μm thick) were cut and mounted on membrane slides (MMI, Rockledge, FL). For tumor blocks, every 4th section was H&E stained and the tumor tissue was marked by a breast pathologist (GMC). For ipsilateral-normal samples, benign glandular areas were selected from blocks that were 3 cm from the tumor tissue. For contralateral-normal samples, benign glandular areas were selected. For both ipsilateral and contralateral samples, glandular tissue was marked on an H&E slide by a pathologist (GMC). For each sample the 6-μm sections mounted on membrane slides were deparaffinized in 3 changes of Xylene and allowed to air dry under vacuum in a desiccator for 30 min prior to the laser-capture microdissection. The unstained sections were oriented for microdissection aided by landmarks defined on the H&E stained slides. Areas to be microdissected were circumscribed using MMI Cell Tools software (Version Celltools-4.4 #261, Rockledge FL). Microdissected tumor, ipsilateral-normal, and contralateral-normal samples from each patient were collected separately onto caps. The collected tissue was placed in 150 μL digestion buffer containing 10 μL proteinase K, kept overnight at 55°C, and stored at −80°C until further processing. DNA was isolated using the manufacturer’s protocol for the BiOstic DNA FFPE Tissue Kit (MoBio).

### DNA methylation profiling

DNA samples were sent to the Molecular Genomics/Methylation Core at the University of Southern California. Bisulfite treatment with the EZ DNA methylation kit (Zymo) was performed and modified DNA was assessed with four PCR reactions to estimate quantity and test for complete bisulfite conversion [8]. DNA samples that passed the PCR quality controls were amplified at 37°C for 20-24 hours after treatment with 0.1N NaOH. The DNA was fragmented at 37°C for 1 hour and subsequently precipitated in 100% 2-propanol at 4°C for 30 minutes followed by centrifugation at 3000xg at 4°C for 20 minutes. Dried DNA-pellets were resuspended in hybridization buffer at 48°C for 1 hour, denatured for 20 minutes at 95°C and loaded onto the Human MethylationEPIC BeadChip, and hybridized at 48°C for 16-24 hours. Unhybridized and non-specific DNA was removed using wash buffers. After a single base extension of the hybridized primers using labeled nucleotides, the BeadChip was stained with Cy-3 and Cy-5 fluorescent dyes and read using the Illumina iScan Reader.

### DNA methylation data processing

Sample intensity data (IDAT) files were processed with the R package *minfi* using the “Funnorm” normalization method on the full dataset. CpGs with a detection *p* value > 1E-06 in more than 5% of samples, CpGs with high frequency SNP(s) in the probe, probes previously described to be potentially cross-hybridizing, and sex-specific probes were filtered. Samples with more than 10% of probes above the detection *p* value or with bisulfite conversion intensity less than 3 standard deviations of the mean were removed.

One ipsilateral-normal sample was removed due to poor sample quality (>10% of probes above the detection *p* value). From an original set of 853,307 measured CpG loci on the Illumina HumanMethylationEPIC array (EPIC), our filtering steps removed 96,564 probes exceeding the detection *p* value limit, 71,478 probes that were SNP associated or cross-hybridizing, and 15,585 sex-specific probes. The final data for analysis resulted in 683,209 CpG loci measured in 25 matched samples from nine women (Table 1) from tumor tissue (*n* = 8), ipsilateral-normal tissue (*n* = 8), and contralateral-normal tissue (*n* = 9).

**Table 1.** Selected subject and tumor characteristics

| Subject | Age | Parity | Carcinoma | ER | PR | HER2 | Size<br>(cm) | Grade | Contralateral<br>Normal<sup>a</sup> | Ipsilateral<br>Normal<sup>a</sup> | Tumor<sup>a</sup> |
| --- | --- | --- | --- | --- | --- | --- | --- | --- | --- | --- | --- |
| 1 | 47 | 2 | Invasive | + | + | + | 1.4 | 2 | 1 | 0 | 1 |
| 2 | 66 | 4 | Invasive | + | - | - | 0.2 | 3 | 1 | 1 | 1 |
| 3 | 61 | 2 | Invasive | + | + | + | 2.5 | 3 | 1 | 1 | 1 |
| 4 | 42 | 2 | In Situ | + | + | N/A | N/A | N/A | 1 | 1 | 1 |
| 5 | 52 | 2 | N/A | N/A | N/A | N/A | N/A | N/A | 1 | 1 | 0 |
| 6 | 49 | 2 | Invasive | + | + | + | 2.1 | 2 | 1 | 1 | 1 |
| 7 | 46 | 4 | Invasive | + | + | N/A | 2.5 | 1 | 1 | 1 | 1 |
| 8 | 50 | 0 | In Situ | + | + | - | N/A | N/A | 1 | 1 | 1 |
| 9 | 32 | 1 | Invasive | + | + | + | 5.1 | 3 | 1 | 1 | 1 |

+ = positive; - = negative; N/A = not available; <sup>a</sup> Number of samples included in the analysis

### Statistical analysis

Unsupervised hierarchical clustering was conducted based on Manhattan distance and using the 10,000 CpG loci that demonstrated the most variability in methylation beta value across all samples. To quantify differential methylation, pairwise comparisons were run on logit-transformed methylation beta values (M-values) between contralateral-normal and tumor tissue, ipsilateral-normal and tumor tissue, and contralateral-normal and ipsilateral-normal tissue. Linear mixed-effect models were performed to model the relation between tissue type (tumor relative to contralateral-normal, tumor relative to ipsilateral-normal, and ipsilateral-normal relative to contralateral-normal, respectively) and CpG methylation M-value, controlling for age and parity and allowing a random effect for each subject. Due to the limited sample size of the study, we utilized nominal significance levels of *p* < 0.01 for subsequent analyses and interpretations.

To classify the genomic context of identified differentially methylated CpG loci, annotation data for the Illumina HumanMethylationEPIC array was used to identify the relation to CpG islands for each probe. The UCSC Genome Browser UCSC_hg19_refGene file was used to define promoters, exons, introns, and intergenic regions and the R package *GenomicRanges* was used to map these regions to all CpG loci on the Illumina HumanMethylationEPIC array. In cases where CpG loci mapped to more than one genomic region, loci were preferentially assigned to promoters, then exons, and then introns such that each locus only had one associated genomic region.

Gene set enrichment analyses of identified differentially methylated CpG loci were conducted using the *gometh* function from the *missMethyl* package in R [9]. In brief, this method assesses enrichment of represented gene ontology (GO) terms among a set of differentially methylated CpG loci relative to a specified background set while adjusting for potential bias introduced by differences in the number of probes that map to each gene leading to altered probability of identifying a differentially methylated locus across different genes.

Cochran-Mantel-Haenszel tests were used to calculate odds ratios (ORs) and *p* values for the enrichment of CpG island contexts, adjusting for Illumina probe type, among CpG loci that were significantly hypomethylated or hypermethylated (*p* < 0.01) in ipsilateral-normal relative to contralateral-normal tissue, compared to all other CpG loci included in the analysis.

To assess possible enrichment of regulatory elements among hypomethylated and hypermethylated (*p* < 0.01) loci, respectively, in ipsilateral-normal relative to contralateral-normal tissue, we utilized Locus Overlap Analysis (LOLA) [10] using Cistrome data for MCF-7 and T47D breast cancer cell lines from the LOLA core database for *hg19*. In brief, LOLA conducts a Fisher’s Exact Test for the enrichment of the identified differentially methylated loci relative to a specified background set (here specified as the 683,209 loci included in the analysis) overlapping with specified regulatory elements.

Methylation status of *Alu* and long interspersed nucleotide element-1 (*LINE-1*) repetitive elements were predicted using the *REMP* package in R [11]. Median value of *Alu* and *LINE-1* were subsequently calculated across all reported regions for each sample. Median *Alu* and *LINE-1* methylation status was then compared between tissue types using a Wilcoxon rank-sum test.

## Results

### Global trends in DNA methylation suggest differences between ipsilateral-normal and contralateral-normal breast tissue

Unsupervised hierarchical clustering using mean methylation beta values of the top 10,000 most variable probes across the three tissue types showed clear separation between tumor and normal (ipsilateral or contralateral) tissue (Figure 1). Among the cluster of normal tissue, some samples clustered by subject (subjects 4, 7, and 8) while some samples appeared to separate by the breast from which they were collected (ipsilateral or contralateral). When assessing the methylation status of *Alu* and long interspersed nucleotide element-1 (*LINE-1*) repetitive elements, there was a statistically significant decrease in median *Alu* element methylation in tumor relative to contralateral-normal tissue (*p* = 0.03) while no statistically significant differences were observed in ipsilateral-normal tissue relative to tumor or contralateral-normal tissue (Figure S1). However, there was visual suggestion of a trend in decreased *Alu* element methylation across contralateral-normal, ipsilateral-normal, and tumor tissue respectively. No statistically significant differences were observed in the methylation status of *LINE-1* elements across tissue types.

**Figure 1.**
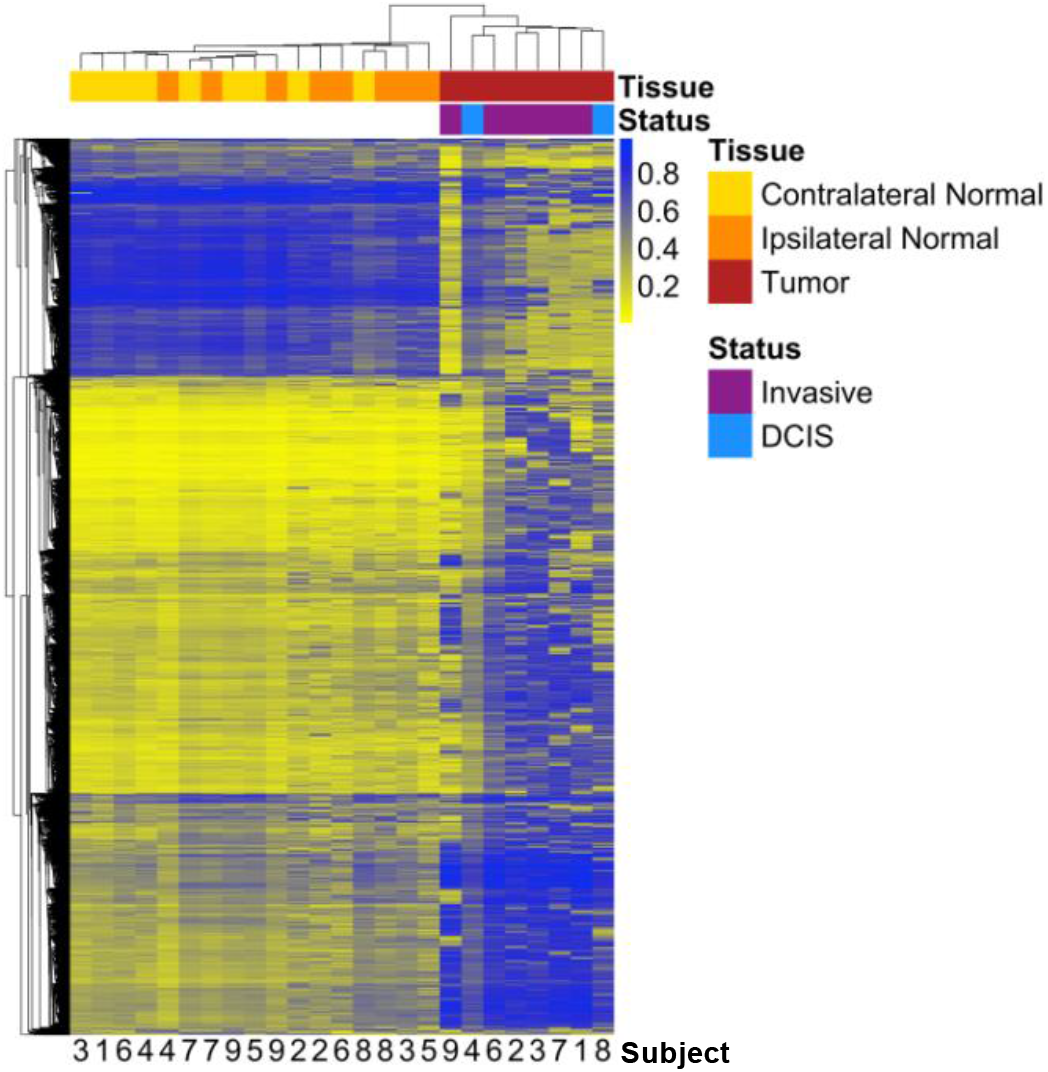
Heatmap with unsupervised hierarchical clustering based on Manhattan distance of tumor, ipsilateral-normal, and contralateral-normal methylation profiles

### Contralateral-normal tissue provides a distinct reference epigenome compared to ipsilateral-normal tissue

At an FDR cutoff of *q* < 0.01, a total of 38,346 CpG loci were identified as differentially methylated in tumor tissue relative to ipsilateral-normal tissue (Figure 2B). However, when comparing tumor tissue to contralateral-normal tissue, 63,271 CpG loci were identified as differentially methylated (Figure 2A), a 65% increase. Furthermore, among those differentially methylated loci, 33,657 (88% of those identified as differentially methylated using ipsilateral-normal as the referent tissue and 53% of those identified as differentially methylated using contralateral-normal as there referent tissue) were identified in both comparisons (Figure 2C) with consistent directionality of association in tumor relative to the respective referent tissue.

**Figure 2.**
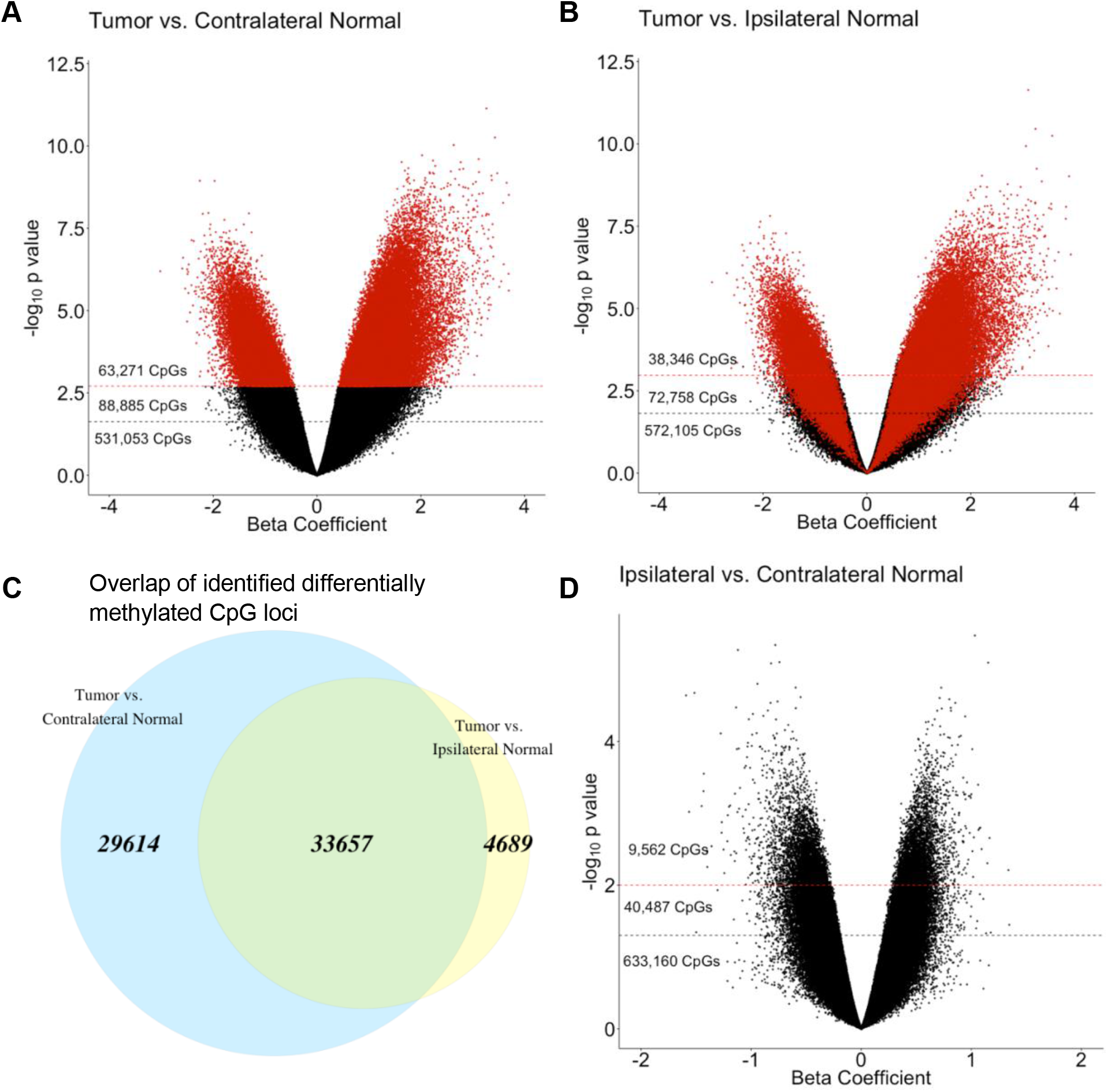
Epigenome-wide association analyses identifying CpG sites that are significantly differentially methylated in (A) tumor relative to contralateral-normal tissue and (B) tumor relative to ipsilateral-normal tissue. The beta coefficient reflects the difference in M-value associated with tissue type, adjusted for subject age and parity. Red dashed lines indicate a significance threshold of q < 0.01 and black dashed lines indicate a significance threshold of q < 0.05. CpGs are colored (red) by sites identified as significant in the tumor versus contralateral-normal tissue (q < 0.01) analysis. (C) Overlap (green) between significantly differentially methylated CpG loci (q < 0.01) in tumor versus contralateral-normal and tumor versus ipsilateral-normal tissue. (D) Epigenome-wide association analysis identifying CpG sites that are significantly differentially methylated in ipsilateral-normal relative to contralateral-normal tissue. Red dashed lines indicate a significance threshold of p < 0.01 and black dashed lines indicate a significance threshold of p < 0.05.

In a sensitivity analysis removing the contralateral normal-sample from the subject whose ipsilateral-normal sample was removed from analysis during QC due to > 10% of probes being above the specified detection *p* value (Subject 1) to allow for matched sample size across comparisons, a modest decrease was observed in the number of loci identified as differentially methylated (Figure S2). However, there remained a 23% increase in the number of differentially methylated loci observed relative to the comparison using ipsilateral-normal tissue as the referent tissue.

To further assess whether the observed increase in differentially methylated CpGs were functionally redundant and reflected more loci from the same genes being identified as differentially methylated, we investigated the overlap in genes and gene sets. The 38,346 differentially methylated CpG loci identified when comparing tumor to ipsilateral-normal tissue mapped to 9,800 unique genes. Conversely, the 63,271 differentially methylated CpG loci identified when using contralateral-normal as the referent tissue mapped to 12,887 genes, 9,283 of which were overlapping with those identified in the tumor relative to ipsilateral-normal comparison.

We used the *gometh* function from the *missMethyl* package in R to conduct gene set enrichment analyses accounting for the bias of the number of probes representing each gene on the EPIC array to assess gene sets that were enriched among the 29,614 CpG loci identified as differentially methylated in tumor relative to contralateral-normal tissue that were not identified when using ipsilateral normal tissue as the referent tissue. These 29,614 were compared to the background set of all 63,271 differentially methylated loci identified between tumor and contralateral-normal tissue to investigate GO terms that may be uniquely implicated in early carcinogenesis. We identified significant enrichment of (*q* < 0.01) 83 GO terms with the most significantly enriched GO terms relating to biological processes such as cell death, metabolic processes, and tissue development (Table S1).

### Hypomethylated loci in ipsilateral-normal relative to contralateral-normal tissue are enriched for transcription factor binding sites implicated in breast carcinogenesis

In analyses comparing ipsilateral-normal to contralateral-normal tissue, 9,562 CpG loci were identified as differentially methylated between the two histologically normal tissue types at a nominal significance threshold of *p* < 0.01 (Figure 2D). Given differences in the biological interpretation of hypomethylation and hypermethylation, respectively, subsequent analyses were stratified by the direction of the identified association.

Among the 9,562 differentially methylated loci, 4,942 were identified as hypomethylated in ipsilateral-normal relative to contralateral-normal tissue (Table S2). These hypomethylated loci demonstrated significant enrichment for CpG sparse open sea regions (OR = 1.77; 95% CI: 1.66, 1.88; *p* = 5.1 × 10^−63^) and significant depletion for all other CpG island associated genomic contexts (Figure 3A).

**Figure 3.**
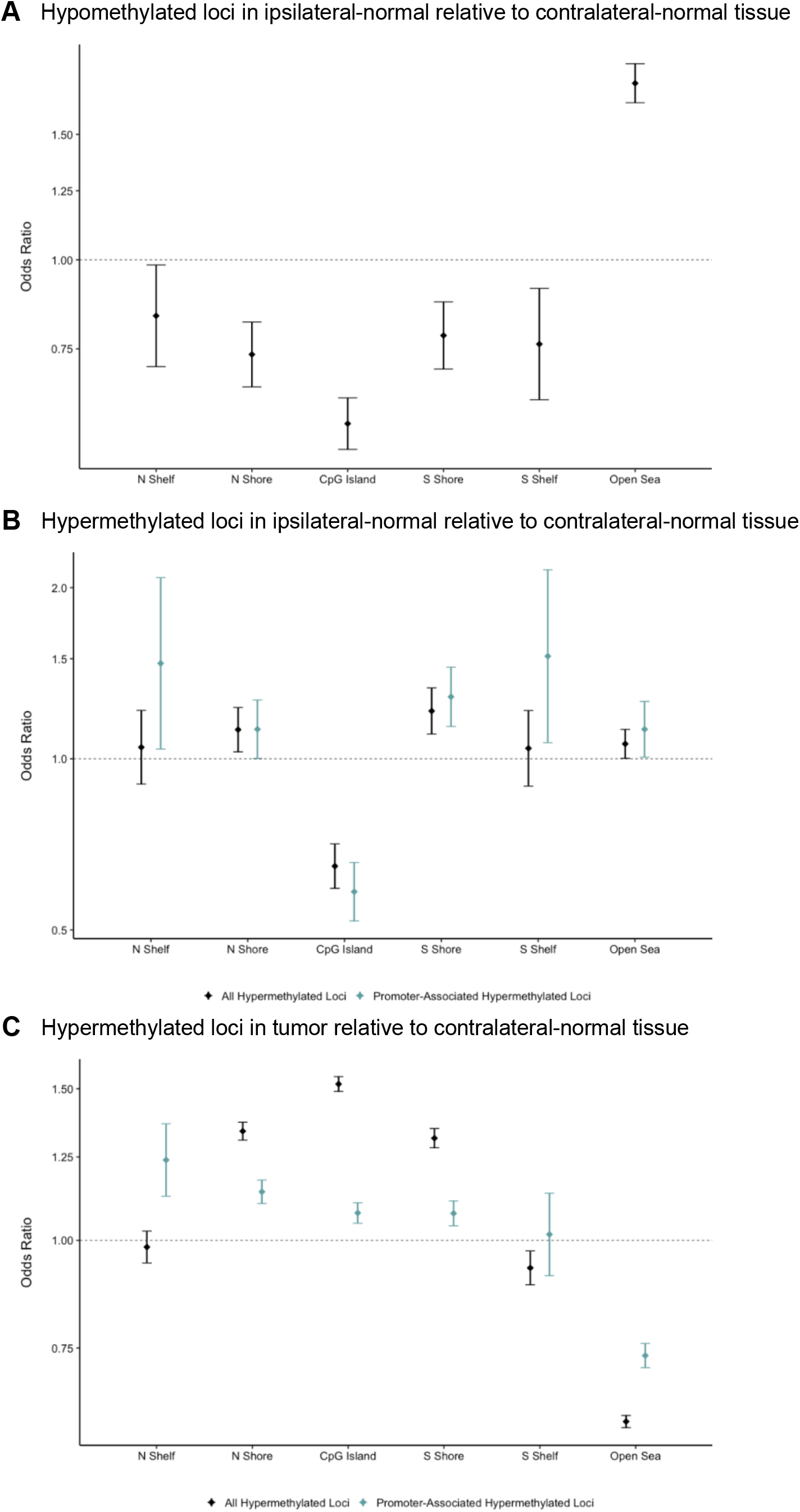
(A) Enrichment/depletion of CpG island contexts from a Cochran-Mantel-Haenszel test comparing identified CpG loci to all other CpG loci included in the analysis, adjusting for Illumina probe type (A) among the 4,942 CpG loci found to be significantly (p < 0.01) hypomethylated in ipsilateral-normal relative to contralateral-normal tissue, (B) among the 4,620 CpG loci found to be significantly (p < 0.01) hypermethylated in ipsilateral-normal relative to contralateral-normal tissue as well as the subset of 1,421 hypermethylated loci annotated to gene promoter regions, (C) among the 71,407CpG loci found to be significantly (p < 0.01) hypermethylated in tumor relative to contralateral-normal tissue as well as the subset of 30,455 hypermethylated loci annotated to gene promoter regions.

To further explore possible regulatory roles of differentially methylated CpG loci in ipsilateral-normal relative to contralateral-normal tissue we next assessed enrichment for transcription factor binding sites (TFBSs), using the Locus Overlap Analysis (LOLA) software tool. Using the LOLA core database for *hg19*, we assessed enrichment for these loci at TFBSs using Cistrome data for the breast cancer cell lines MCF-7 and T47D. Comparing the identified hypomethylated loci (*p* < 0.01) in ipsilateral-normal relative to contralateral-normal tissue to the background set of all 683,209 loci included in our epigenome-wide association study, we identified statistically significant enrichment of TFBSs for c-Fos, ESR1, FoxA1, GATA3, RAD21, RARA, RARG, and SRC-3 using MCF-7 data and ESR1, FoxA1, and PGR using T47D data (all *q* < 0.05, Figure 4). This suggests that cellular programs regulated by transcription factors known to be dysregulated in invasive breast cancer also contribute to early events in breast carcinogenesis.

**Figure 4.**
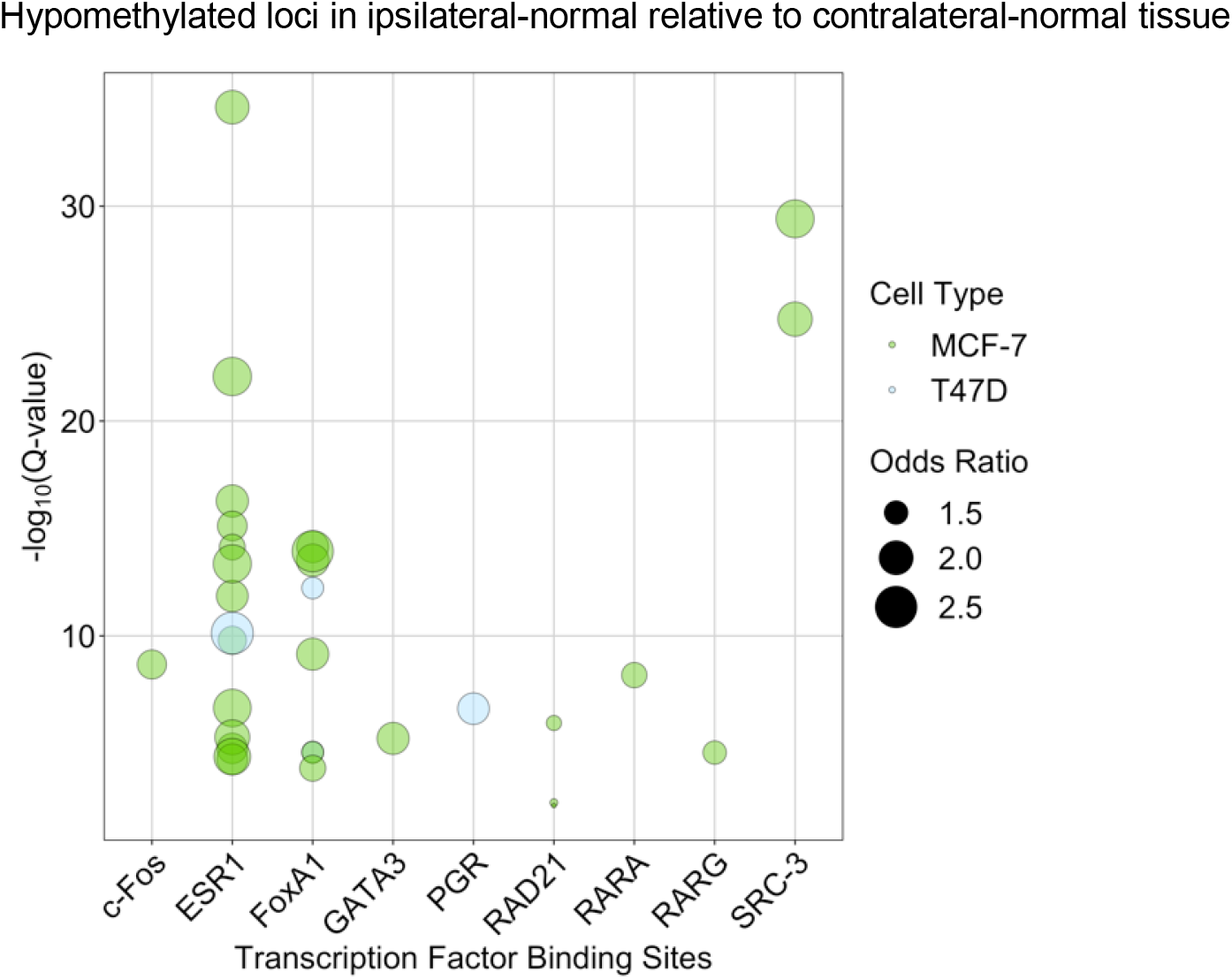
Enrichment of transcription factor binding sites in MCF-7 and T47D cell lines as assessed in LOLA using Cistrome data among the 4,942 CpG loci found to be significantly (p < 0.01) hypomethylated in ipsilateral-normal relative to contralateral-normal tissue.

### Hypermethylated loci in ipsilateral-normal relative to contralateral-normal tissue are depleted for CpG islands but enriched for adjacent shore regions

While the 4,620 loci identified as hypermethylated in ipsilateral-normal relative to contralateral-normal tissue (Table S3) also demonstrated modest enrichment for open sea regions (OR = 1.06; 95% CI: 1.00, 1.13, *p* = 0.044), they also demonstrated significant depletion for CpG island regions (OR = 0.65; 95% CI: 0.59, 0.71, *p* = 6.8 × 10^−20^) but significant enrichment for CpG island north shore region (OR = 1.13; 95% CI: 1.03, 1.23; *p* = 0.010) and south shore region (OR = 1.21; 95% CI: 1.11, 1.33; *p* = 5.7 × 10^−5^; Figure 3B). As not all CpG islands track to promoter regions, we further assessed enrichment of hypermethylated loci in ipsilateral-normal breast tissue restricting only to promoter CpG islands. This analysis demonstrated further depletion for CpG island regions (OR = 0.58; 95% CI: 0.52, 0.66; *p* = 8.2 × 10^−18^) and further enrichment for CpG island south shore regions (OR = 1.29; 95% CI: 1.14, 1.45, *p* = 4.8 × 10^−5^; Figure 3B). In contrast, when assessing the genomic context distribution among the 71,407 hypermethylated loci in tumor relative to contralateral-normal tissue (*p* < 0.01), strong enrichment for both CpG island regions (OR = 1.52; 95% CI: 1.49, 1.55; p = 1.9 × 10^−319^) and bordering CpG island north shore (OR = 1.34; 95% CI: 1.31, 1.37; 1.7 × 10^−110^) and south shore regions was observed (OR = 1.31; 95% CI: 1.28, 1.35, p = 5.2 × 10^−85^; Figure 3C). Using the LOLA core database for *hg19*, no statistically significant enrichments for TFBSs were identified among the hypermethylated loci in ipsilateral-normal relative to contralateral-normal tissue.

## Discussion

Our findings suggest that within the same patient, histologically normal breast tissue from the same breast as a tumor is epigenetically distinct from histologically normal tissue from the opposite breast. Furthermore, histologically normal tissue collected from the breast without the tumor appears to be more epigenetically distinct from tumor tissue than histologically normal tissue collected from the same breast as the tumor. This is suggestive of a field cancerization effect of the tumor on surrounding histologically normal tissue.

While differences between ipsilateral-normal and contralateral-normal tissue were modest and did not achieve statistical significance at an FDR threshold of *q* < 0.05, our study was limited by small sample size. Furthermore, differentially methylated CpG loci at a nominal significance threshold of *p* < 0.01 were significantly enriched for regions of functional significance including CpG island shore regions and tissue specific regulatory regions. This suggests that these differences may have important implications for the regulation of gene expression between the two tissue types.

Furthermore, enrichment of CpG island shore regions but depletion of CpG island regions among hypermethylated loci in ipsilateral-normal relative to contralateral-normal tissue, with greater magnitudes of enrichment and depletion observed when restricting to gene promoter regions, suggests that the earliest molecular alterations in carcinogenesis might appear as hypermethylation of CpG island shore regions and that these early alterations may also be present in histologically normal tissue surrounding the tumor. These findings are further supported by existing work demonstrating methylation encroachment at the borders of CpG islands in breast cancer and prostate cancer cells [12], suggesting that hypermethylation of promoter regions over the course of carcinogenesis might begin at CpG island shores and progress into the islands.

Histologically normal tissue in the breast harboring tumor also shows evidence of early molecular alterations that may be implicated in carcinogenesis among loci that are hypomethylated in this tissue relative to contralateral-normal tissue. These loci demonstrate enrichment for binding sites for transcription factors previously implicated in breast cancer, suggesting that hypomethylation of these loci may play a role in transcriptional regulation.

We did not identify significant differences between the methylation status of *Alu* and *LINE-1* repetitive elements across tissue types. While previous work has identified hypomethylation of these repetitive elements over the course of carcinogenesis [13], these findings were subtype-specific. Therefore, given our limited sample size, it is unclear whether such associations might be observed given the lack of power to stratify analyses by molecular subtype.

Observed differences have important implications for molecular research in which histologically normal tissue adjacent to a tumor is used as a comparison group when evaluating for molecular alterations in tumor tissue. Given that our study collected ipsilateral-normal tissue at a distance of 3 cm from the tumor, 1 cm more distal from the tumor than the TCGA specified 2 cm minimum distance for the collection of adjacent normal tissue, this is suggestive that adjacent normal tissue may not be an adequate referent tissue. The observed effect of the tumor/tumor microenvironment on adjacent normal tissue may attenuate results when adjacent normal tissue is used as a comparison. In such cases, research may fail to identify significant differences at certain loci between tumor tissue and healthy breast tissue within the same individual due to an unquantified field cancerization effect on the referent tissue. Furthermore, those loci may reflect the earliest alterations to DNA methylation in breast carcinogenesis and, therefore, failing to identify alterations at these loci may limit our understanding of early carcinogenesis. Our findings suggest that additional research is needed to identify molecular differences between breast tumors and normal breast tissue that previous research utilizing histologically normal adjacent tissue may have been unable to capture.

## Conclusions

Our results indicate that histologically normal tissue collected from the same breast as the tumor harbors field cancerization and is not an ideal referent tissue for identifying breast tumor DNA methylation alterations, particularly early events in carcinogenesis. Furthermore, our results suggest that early hypermethylation events in breast carcinogenesis are more likely to occur in the regions immediately surrounding CpG islands than CpG islands. This work offers an opportunity to focus investigations of early DNA methylation alterations in breast carcinogenesis and potentially develop epigenetic biomarkers of disease risk.

## Data Availability

The datasets generated and analyzed during the current study are available in the Gene Expression Omnibus under the accession number GSE133985 (https://www.ncbi.nlm.nih.gov/geo/).

## Abbreviations

DCIS: ductal carcinoma *in situ*
GO: gene ontology; OR: odds ratio
CI: confidence interval
TFBS: transcription factor binding site
LOLA: Locus Overlap Analysis

## Declarations

## Acknowledgements

The authors thank all the women who donated tissue to this study.

## Authors’ contributions

MEM conceived and designed the analytical approach, performed statistical analyses, interpreted the results, and wrote and revised the manuscript. AJT conceived and designed the analytical approach, performed statistical analyses, interpreted the results, and wrote and revised the manuscript. LAS designed the analytical approach, interpreted the results, and revised the manuscript. OMW designed the analytical approach, interpreted the results, and revised the manuscript. CM assisted with microdissection, isolated DNA, interpreted the results, and revised the manuscript. KJG sectioned tissue, performed microdissections, interpreted results, and revised the manuscript. SSS conceived and designed the approach, interpreted the results, and revised the manuscript. GMC reviewed normal and tumor tissue, marked areas for microdissection, interpreted the results, and revised the manuscript. RMJ reviewed normal and tumor tissue, interpreted the results, and revised the manuscript. CNO conceived and designed the approach, interpreted the results, and revised the manuscript. BCC conceived and designed the approach, oversaw project development, interpreted the results, and revised the manuscript. KFA conceived and designed the approach, oversaw project development, interpreted the results, and revised the manuscript. All authors read and approved the final manuscript.

## Ethics approval and consent to participate

Written informed consent was obtained from all subjects. Approval for this research was obtained from the Baystate Health Institutional Review Board (protocol number 568088).

## Consent for publication

Not applicable.

## References

1. Siegel RL, Miller KD, Jemal A. Cancer statistics, 2019. CA Cancer J Clin. American Cancer Society; 2019;69:7–34.

2. Koboldt DC, Fulton RS, McLellan MD, Schmidt H, Kalicki-Veizer J, McMichael JF, et al. Comprehensive molecular portraits of human breast tumours. Nature. Nature Publishing Group; 2012;490:61–70.

3. Christensen BC, Kelsey KT, Zheng S, Houseman EA, Marsit CJ, Wrensch MR, et al. Breast cancer DNA methylation profiles are associated with tumor size and alcohol and folate intake. PLoS Genet. United States; 2010;6:e1001043.

4. McPherson K, Steel CM, Dixon JM. Breast cancer—epidemiology, risk factors, and genetics. BMJ Br Med J. British Medical Journal; 2000;321:624–8.

5. Johnson KC, Koestler DC, Fleischer T, Chen P, Jenson EG, Marotti JD, et al. DNA methylation in ductal carcinoma in situ related with future development of invasive breast cancer. Clin Epigenetics. BioMed Central; 2015;7:75.

6. Fleischer T, Frigessi A, Johnson KC, Edvardsen H, Touleimat N, Klajic J, et al. Genome-wide DNA methylation profiles in progression to in situand invasive carcinoma of the breast with impact on gene transcription and prognosis. Genome Biol. BioMed Central; 2014;15:435.

7. Teschendorff AE, Gao Y, Jones A, Ruebner M, Beckmann MW, Wachter DL, et al. DNA methylation outliers in normal breast tissue identify field defects that are enriched in cancer. Nat Commun. 2016;7.

8. Campan M, Weisenberger DJ, Trinh B, Laird PW. MethyLight. Humana Press; 2009. p. 325–37.

9. Phipson B, Maksimovic J, Oshlack A. missMethyl: an R package for analyzing data from Illumina’s HumanMethylation450 platform. Bioinformatics. 2015;32:btv560.

10. Sheffield NC, Bock C. LOLA: enrichment analysis for genomic region sets and regulatory elements in R and Bioconductor. Bioinformatics. Oxford University Press; 2016;32:587–9.

11. Zheng Y, Joyce BT, Liu L, Zhang Z, Kibbe WA, Zhang W, et al. Prediction of genome-wide DNA methylation in repetitive elements. Nucleic Acids Res. Oxford University Press; 2017;45:8697–711.

12. Skvortsova K, Masle-Farquhar E, Luu P-L, Song JZ, Qu W, Zotenko E, et al. DNA Hypermethylation Encroachment at CpG Island Borders in Cancer Is Predisposed by H3K4 Monomethylation Patterns. Cancer Cell. Elsevier; 2019;35:297–314.e8.

13. Park SY, Seo AN, Jung HY, Gwak JM, Jung N, Cho N-Y, et al. Alu and LINE-1 Hypomethylation Is Associated with HER2 Enriched Subtype of Breast Cancer. El-Maarri O, editor. PLoS One. Public Library of Science; 2014;9:e100429.

